# 3D fusion between SPECT myocardial perfusion imaging and invasive coronary angiography to guide the treatment for patients with stable CAD

**DOI:** 10.1101/2023.09.18.23295731

**Authors:** Zhihui Xu, Saurabh Malhotra, Chen Zhao, Jingfeng Jiang, Aviral Vij, Zekang Ye, Rui Hua, Chunjian Li, Cheng Wang, Weihua Zhou

## Abstract

**Objective:** We evaluated the value of three-dimensional (3D) fusion of SPECT myocardial perfusion imaging (MPI) with invasive coronary angiography (ICA) to guide coronary revascularization for patients with stable coronary artery disease (CAD).

**Methods:** A retrospective observational study of 621 patients who underwent SPECT MPI and ICA was conducted. Based on the location of perfusion deficit on SPECT MPI and stenosis on ICA, patients were classified into matched, unmatched, or normal groups via the fusion or side-by-side analysis. The treatments recommended by the fusion or side-by-side analysis were compared with those that the patients actually received. The treatment was defined as concordant if there was revascularization in concordance with the recommendation by the fusion or side-by-side analysis or if patient did not require revascularization; otherwise, it is classified as discordant. Major adverse cardiac events (MACE) were defined as all-cause and cardiac death, myocardial infarction, unstable angina requiring hospitalization or ICA, or unplanned revascularization.

**Results:** Over a five-year follow-up, 15.9% of patients experienced MACE. The MACE rates in the fusion and side-by-side groups were 19.8% and 24.4% for matched findings, 14.4% and 14.0% for unmatched findings, and 7.2% and 8.2% for normal findings, respectively (P<0.01). Among the 366 patients with at least one vessel stenosis of >50%, those who received the treatment concordant with fusion had significantly better outcomes compared to those who did not (16.8% vs 27.0%, P<0.05), particularly in the sub-group with intermediate stenosis (stenosis: 50-80%) (10.8% vs 26.5%, P<0.01). The treatment concordant with fusion is an independent protective factor against MACE (HR:0.48, CI:0.28-0.83 P<0.01) while the treatment concordant with the side-by-side analysis is not. The concordant group showed a significantly higher lesion vessel rate in the left circumflex artery (LCX) (31.2% vs. 14.3%, P<0.01) compared to the discordant group as classified by fusion.

**Conclusions:** 3D fusion before coronary revascularization can guide the treatment to improve outcomes among patients with known or suspected CAD, particularly those with intermediate stenosis.

## Introduction

Invasive coronary angiography (ICA) is considered as the gold standard for diagnosing and treating coronary artery disease (CAD). However, the large International Study of Comparative Health Effectiveness with Medical and Invasive Approaches (ISCHEMIA) study showed that in patients with stable CAD an initial revascularization strategy did not reduce the risk of ischemic cardiovascular events or overall mortality, compared to a conservative strategy over a 3.2-year follow-up period [1]. The results of this trial has encouraged use of aggressive medical therapy and risk stratification using noninvasive stress modalities for patients with stable ischemic heart disease. There is strong evidence supporting the use of noninvasive testing as the initial approach in managing patients with stable CAD [2]. Myocardial perfusion imaging (MPI) using single photon emission computed tomography (SPECT) is a well-established non-invasive method that has been widely used to assess the functional significance of coronary stenosis [3]. However, SPECT MPI has limitations, such as low sensitivity, challenges in multivessel CAD leading to balanced ischemia, and difficulties in correlating findings with specific lesioned vessels [4].

Fusion cardiac imaging, utilizing a combination of SPECT with coronary computed tomography angiography (CTA), enables the depiction of both myocardial functional and vascular anatomical abnormalities in a single examination [5]. A SPECT-CTA fusion study has demonstrated that the matched finding of anatomical lesion and perfusion deficit is associated with a high cardiac event rate, and it is a good predictor of poor prognosis [6]. However, there are still concerns about the appropriate selection of patients for this integrated examination to achieve optimal clinical effectiveness while minimizing costs and radiation exposure. Moreover, it is uncertain whether the SPECT-CTA fusion will amplify deficiency limitations such as motion artifacts for CTA and balanced ischemia in SPECT.

We have developed a 3D fusion approach that combined SPECT and ICA and analyzed its clinical utility based on per-vessel analysis [7]. Compared with the side-by-side analysis, our SPECT-ICA fusion approach showed a lower number of segments being qualified with equivocal coronary luminal stenosis and an improvement in the revascularization ratio of the matched group [8]. The aim of this study is to evaluate the role of the fusion in guiding treatment decisions for patients with known or suspected CAD.

## MATERIALS AND METHODS

### Study Population

This is a retrospective observational study that reclassifies patients using both the SPECT-ICA fusion and side-by-side analysis, and compares their outcomes, specifically all-cause mortality and cardiac outcomes.

For each patient with suspected CAD, SPECT was acquired before ICA, with a time interval of 9.62 ± 19.4 days between the two procedures (all <3 months). The exclusion criteria were: 1) previous bypass graft lesion or stent implantation, 2) previous myocardial infarction, 3) dilated cardiomyopathy (DCM), 4) hypertrophic cardiomyopathy (HCM), 5) congenital heart disease (CHD). This study complied with the Helsinki declaration and local regulations and was approved by the ethics committee of the First Affiliated Hospital of Nanjing Medical University.

### Image Processing of SPECT MPI

SPECT myocardial perfusion images were obtained using a 2-day stress/rest MPI protocol in all patients with suspected CAD. Stress and rest SPECT images were acquired with a weight-adjusted dose of 400-600 MBq of 99mTc-sestamibi. A dual-headed gamma camera (CardioMD, Philips Medical Systems, Milpitas, California) was used for SPECT imaging, with an energy window set at 20% around the 140-keV photon peak. Transverse tomograms were reconstructed and short-axis, vertical, and horizontal long-axis tomograms were produced. Two experienced nuclear medicine physicians, who were blinded to other imaging data, reported the results in consensus. Images were interpreted based on a 17-segment model with a five-point scoring system (0 = normal to 4 = absence of tracer uptake). The percentage summed difference score (SDS) (SD%), representing the percentage measusure of left ventricular perfusion, is calculated by dividing the SDS by a number corresponding to the SDS value indicating a deficit of 4 in every segment (68 points for 17 segments) [9]. According to current clinical recommendations, perfusion deficit in the anterior and septal wall were allocated to the region of the left anterior descending coronary artery (LAD), deficit in the lateral wall to the region of the left circumflex coronary artery (LCX), and deficit in the inferior wall to the region of the right coronary artery (RCA).

## Image Processing of ICA

ICA was performed using standard percutaneous techniques and revascularization was performed as part of coronary angiography, with stenting performed immediately after ICA at the operator’s discretion. Two experienced interventional cardiologists interpreted all invasive studies and assessed the severity of stenosis, with results reported by consensus. Lesions in the left main coronary were recorded as being in the LM, while diagonal lesions were considered as being in the LAD and obtuse marginal artery lesions were considered as being in the LCX. Lesions in the posterior descending branch were recorded as being in the RCA. The coronary artery tree was subdivided according to the American Heart Association guidelines, and each vessel segment was visually evaluated with vessel border delineation on at least 2 different projections. All coronary arteries larger than 1.5 mm in diameter were analyzed, including distal vessels and those with complete total occlusions.

### Image Fusion between SPECT MPI and ICA

Image fusion between SPECT MPI and ICA was carried out using a previously published approach by a trained operator [7]. A deep learning model was utilized to extract coronary arteries automatically from ICA images, and manual corrections were made when the arteries were not accurately extracted [10, 11]. Once the arterial extraction was completed, 3D arterial anatomy was reconstructed from two projection views and fused with the 3D left-ventricular epicardial surface from SPECT MPI. Functional information such as myocardial perfusion was displayed on the LV surface for data analysis. The fused model could be viewed from various angles with the help of 3D operations like rotation, shift, and scale.

### Data Interpretation by the Side-by-side Analysis and 3D Fusion

In this study, the relationship between the perfusion deficit and coronary artery stenosis was classified into three categories: matched group, unmatched group, and normal group. The matched group was defined as a SPECT MPI deficit in a territory subtended by a stenotic coronary artery, while the unmatched group consisted of those in whom abnormality on SPECT MPI and stenosis on ICA were in unrelated territories. The normal group consisted of patients with normal coronary angiography findings or any luminal narrowing <=50% and no fixed or reversible deficit in SPECT. Treatments that the patients received were compared, on a per patient basis, between those with the guidance by the side-by-side analysis and guidance by fusion results (fusion of SPECT MPI and ICA findings). Concordance was defined as 1) patients in the matched group receiving revascularization; or 2) patients in the normal group who did not receive revascularization. Otherwise, they were regarded as being discordant.

### Patient Follow-up

Follow-up data were collected through standardized telephone interviews and hospital records. The primary outcome measure was the occurrence of major adverse cardiac events (MACE), which included all-cause and cardiac death, myocardial infarction, unstable angina requiring hospitalization or ICA performed more than 90 days after the initial SPECT and ICA test, and unplanned revascularization.

### Statistical Analysis

Statistical analysis was conducted using SPSS 22.0 (IBM, Armonk, NY). Mean ± SD was used to represent numeric variables, while percentages or numbers were used for categorical variables. Continuous variables were compared using t-tests for normally distributed continuous variables. Categorical variables were compared using either the chi-square test (χ2) or Fisher’s exact test. McNemar’s test and Kappa coefficient were employed to evaluate differences (agreement) in perfusion and vessel lesion classification between the side-by-side and fusion analyses. Cumulative event-free survival curves were obtained using the Kaplan-Meier method, and the log-rank test was used to compare the survival curves. Univariate and multivariate Cox regression analysis were conducted to evaluate the impact of revascularizations guided by fusion or side-by-side on MACE while controlling for other clinical characteristics. Variables included in the models were age, male sex, more than three risk factors (i.e., hypertension, diabetes, smoking, drinking, positive family history for CAD), perfusion deficit at SPECT MPI, stenosis of 80% or greater, revascularization, and fusion concordance finding. The regression results were expressed as hazard ratios (HRs) with the respective 95% confidence interval. A significance level of P < 0.05 was considered statistically significant.

## Results

### Patient Population

Out of the total 1104 patients, 149 (13.5%) were lost to follow-up. Additionally, 32 (2.9%) patients were excluded due to significant valve disease, 53 (4.8%) patients were excluded as they had HCM or DCM, 145 (13.1%) patients were excluded due to the loss of image and tag information, and 104 (9.4%) patients were excluded due to previous MI, stents transplantation or coronary artery bypass surgery (CABG). A total of 621 patients with a mean follow-up of 5 years were enrolled and analyzed using the side-by-side and fusion methods (Fig.1).

**Figure 1.**
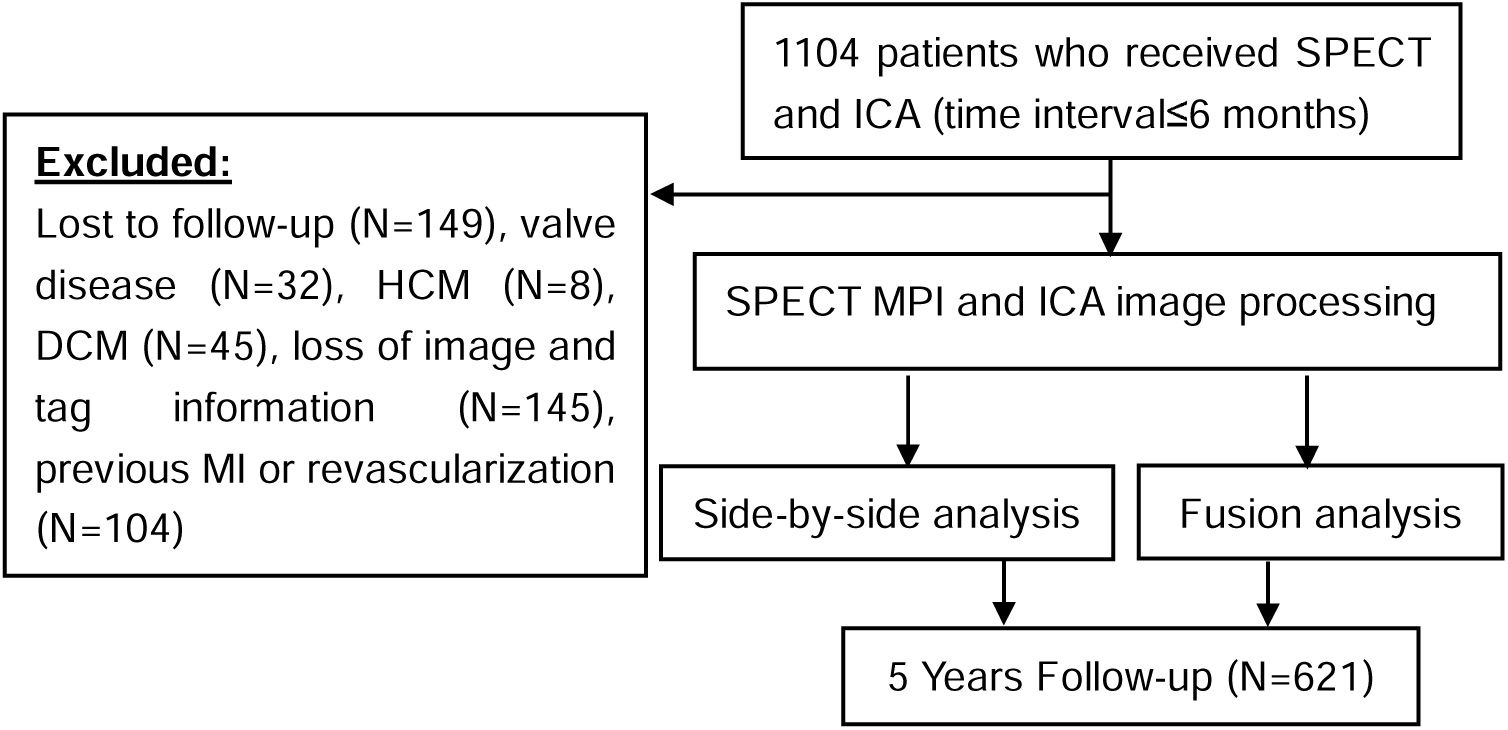
Study flowchart showing patient enrollment.

The average age of the cohort was 61.5 ± 10.5 years, with a mean body mass index of 24.5 ± 3.79. The main presenting symptoms were chest pain (72.1%) and dyspnea (19.3%). The geometric means of summed stress score (SSS), summed rest score (SRS), SDS were 3.48 ± 0.36, 3.65 ± 0.48 and 2.46 ± 0.34, respectively. Table 1 summarizes the patient cohort’s general clinical, angiographic, SPECT MPI, and echocardiography characteristics.

**Table.1.**
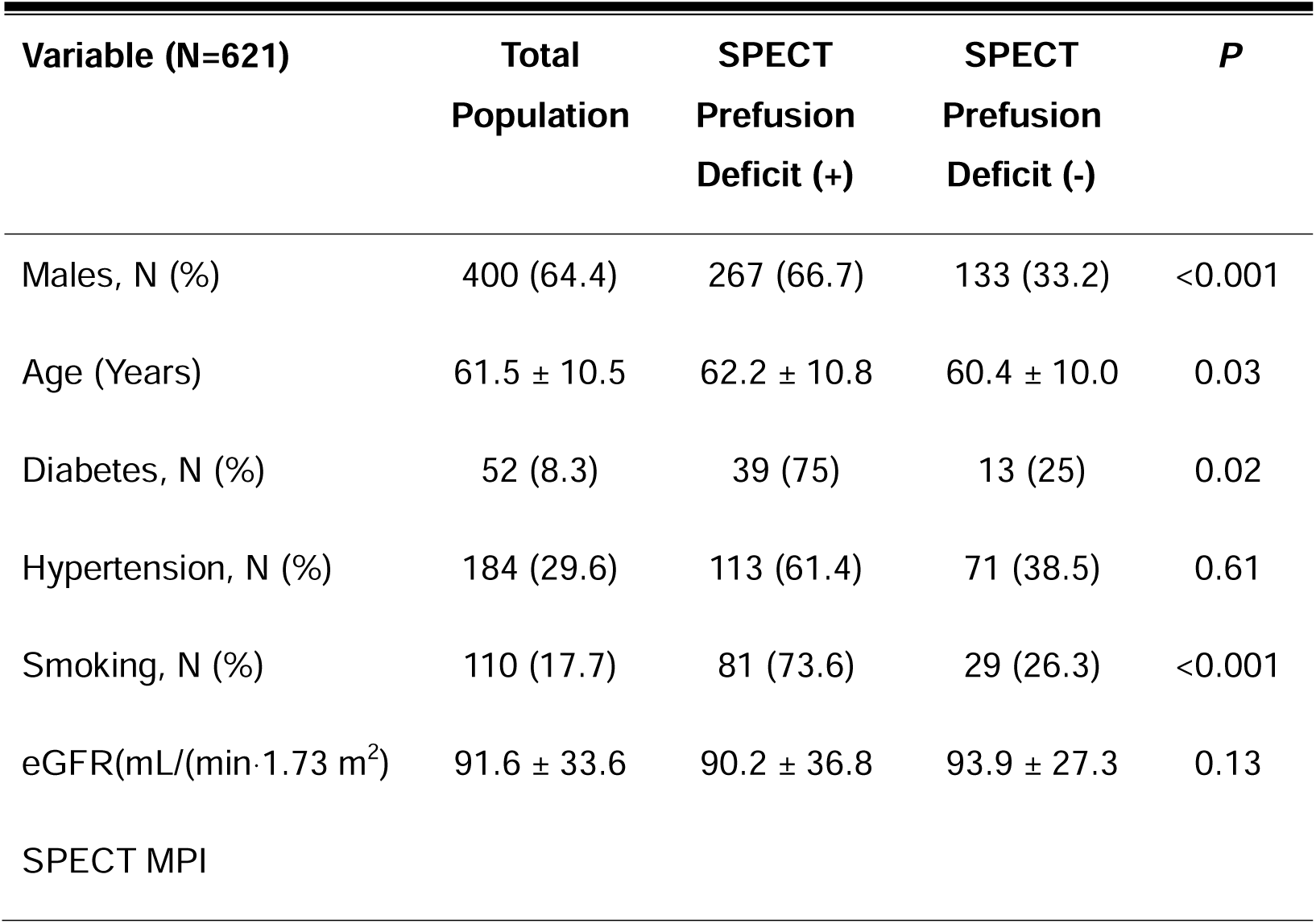
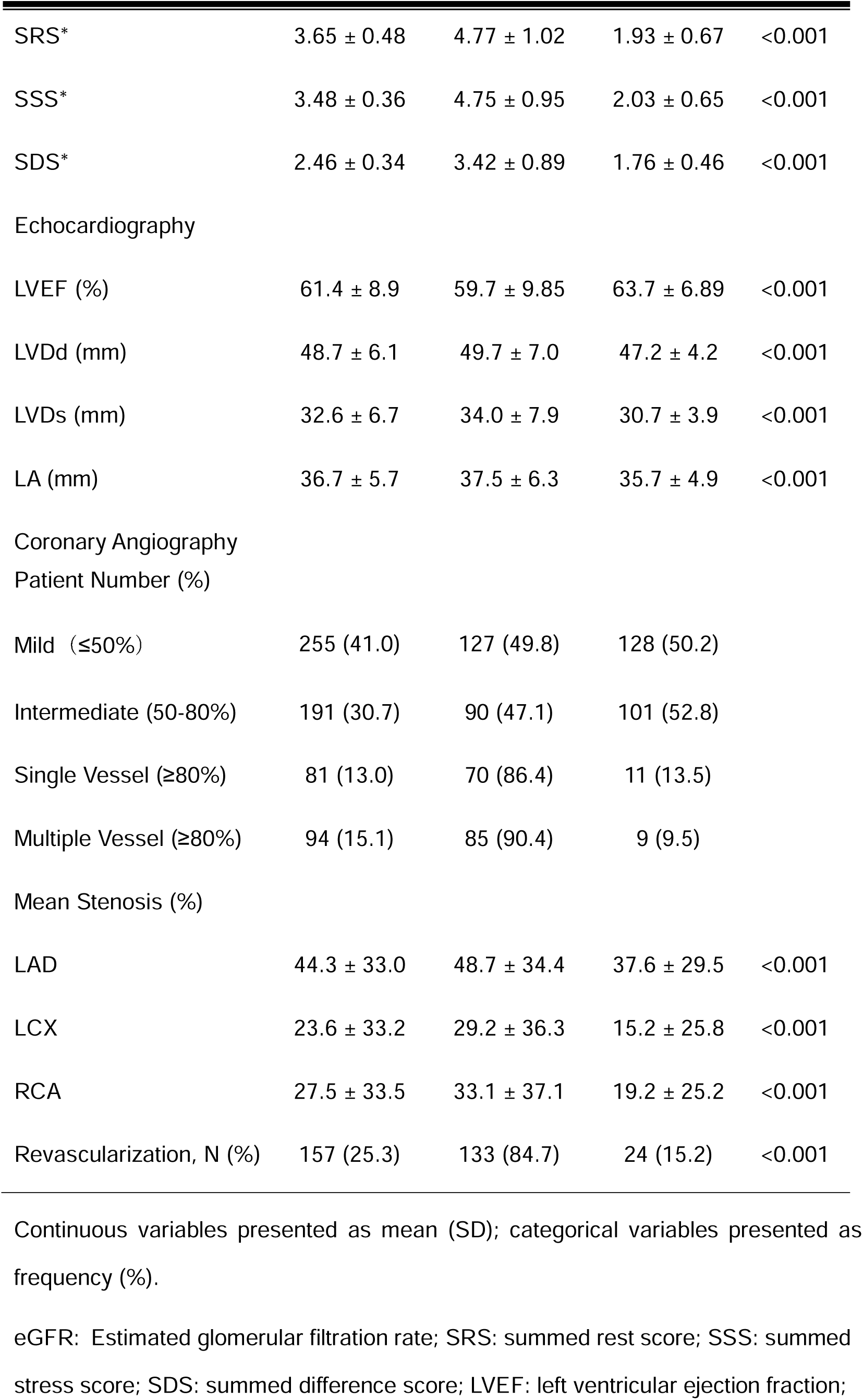
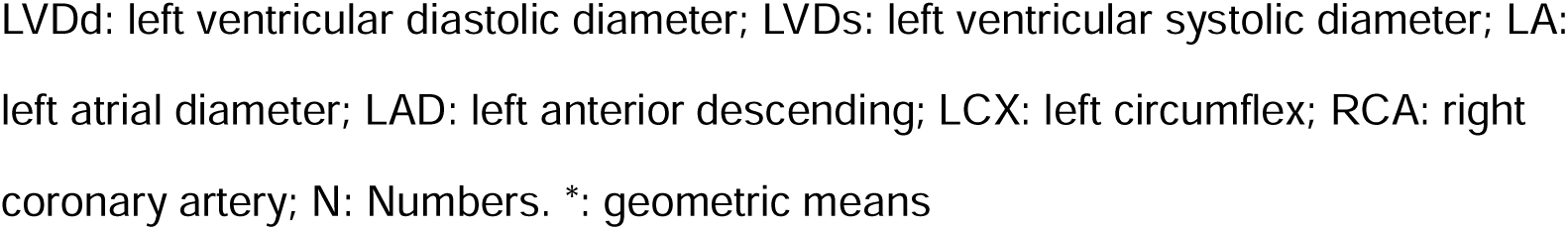
Baseline characteristics of the study population.

Patients with a perfusion deficit were generally more likely to be older, diabetes, smoking, males. The population with positive perfusion deficit exhibited lower ejection fraction (EF) and higher stenosis in the LAD, LCX, and RCA (P<0.05).

### Follow-up on MACE

Over a five-year follow-up, 99 patients experienced the primary endpoints, including all-cause (N=26), myocardial infarction (N=14), unstable angina requiring hospitalization or ICA (N=39), and unplanned revascularization (N=20). On univariate analysis, the presence of a perfusion deficit was a significant predictor of MACE (HR: 1.89, 95% CI 1.21–2.93), along with vessel stenosis>50% (HR:2.61, 95% CI 1.62-4.19) and revascularization (HR: 1.65, 95% CI 1.09–2.49). The predictors remained statistically significant in multivariable analysis as well (perfusion deficit [HR: 1.64, 95% CI 1.02–2.64] and vessel stenosis>50% [HR: 2.43, 95% CI 1.45–4.09]). SPECT perfusion deficit and vessel stenosis>50% were associated with an increased risk of the primary endpoint at over 5 years. There was no significant association between other clinical characteristics and MACE (Table 2).

**Table 2.** Predictors of events at univariate and multivariate analyses (N=621)

| Predictors | MACE |  |  |  |
| --- | --- | --- | --- | --- |
|  | Univariate HR<br>(95% CI) | P-value | Multivariate HR<br>(95% CI) | P-value |
| Male gender | 1.00 (0.67-1.51) | 0.98 | 0.89 (0.58-1.37) | 0.60 |
| Age (≥65) | 1.47 (0.99-2.19) | 0.05 | 1.27 (0.85-1.90) | 0.24 |
| ≥3 risk factors | 0.96 (0.64-1.45) | 0.87 | 0.84 (0.56-1.27) | 0.41 |
| Perfusion deficit | 1.89 (1.21-2.93) | **0.01** | 1.64 (1.02-2.64) | **0.04** |
| Stenosis>50% | 2.61 (1.62-4.19) | **<0.001** | 2.43 (1.45-4.09) | **<0.001** |
| Revascularization | 1.65 (1.09-2.49) | **0.02** | 0.97 (0.61-1.56) | 0.92 |

MACE, major adverse cardiac events; HR, hazard ratio; CI, confidence interval; SSS: summed stress score

### ICA, SPECT, and fusion findings

The study analyzed a total of 40,572 coronary segments from 2,898 coronary arteries. Of these, 255 patients with a stenosis of <=50% were considered normal. Intermediate stenosis (50-80%) was found in 191 patients, while 81 patients had only one vessel disease with above 80% stenosis. Ninety-four patients had at least 2 with > 80% stenosis. Among luminal stenosis >50%, 230 segments were in the LAD, 118 in the LCX, and 131 in the RCA. SPECT imaging revealed abnormal perfusion in 372 patients. Of these perfusion deficit, 164 were in the anterior, 46 in the septal wall, 87 in the inferior wall, and 75 in the posterior and lateral wall. 249 patients did not show any perfusion abnormalities on SPECT.

In this study, all patients were divided into matched, unmatched, or normal groups using the fusion and side-by-side analysis respectively. Compared to the side-by-side analysis, the patient number by the fusion analysis was significantly higher in the matched group (34.9% vs. 55.2%), lower in the unmatched (35.6% vs. 24.6%) and normal groups (29.5% vs. 20.1%). The revascularization ratio reclassified by the fusion analysis was higher in the matched group (21.1% vs. 19.8%) and lower in the unmatched group (3.1% vs. 5.0%), compared to the side-by-side analysis (Table 3).

**Table 3.** Patients number and revascularization ratio by the fusion and side-by-side analysis.

|  | Side-by-side analysis<br>(Revascularization ratio) | SPECT-ICA fusion<br>(Revascularization ratio) |
| --- | --- | --- |
| Normal | 183 (0.5%) | 125 (1.1%) |
| Matched | 217 (19.8%) | 343 (21.1%) |
| Unmatched | 221 (5.0%) | 153 (3.1%) |

### Outcomes

On Kaplan-Meier survival analysis, patients in the normal group classified by the fusion analysis had the best survival(7.2%), while the outcome was progressively worse in the unmatched(14.4%)and matched(19.8%)groups (P<0.01) (Fig.2 A). This result is similar among patients classified by the side-by-side analysis (Fig.2 B), normal findings had excellent event-free survival (8.2%), and the outcome was progressively worse in patients with unmatched (14.0%) and matched findings (24.4%) (P<0.01).

**Figure 2.**
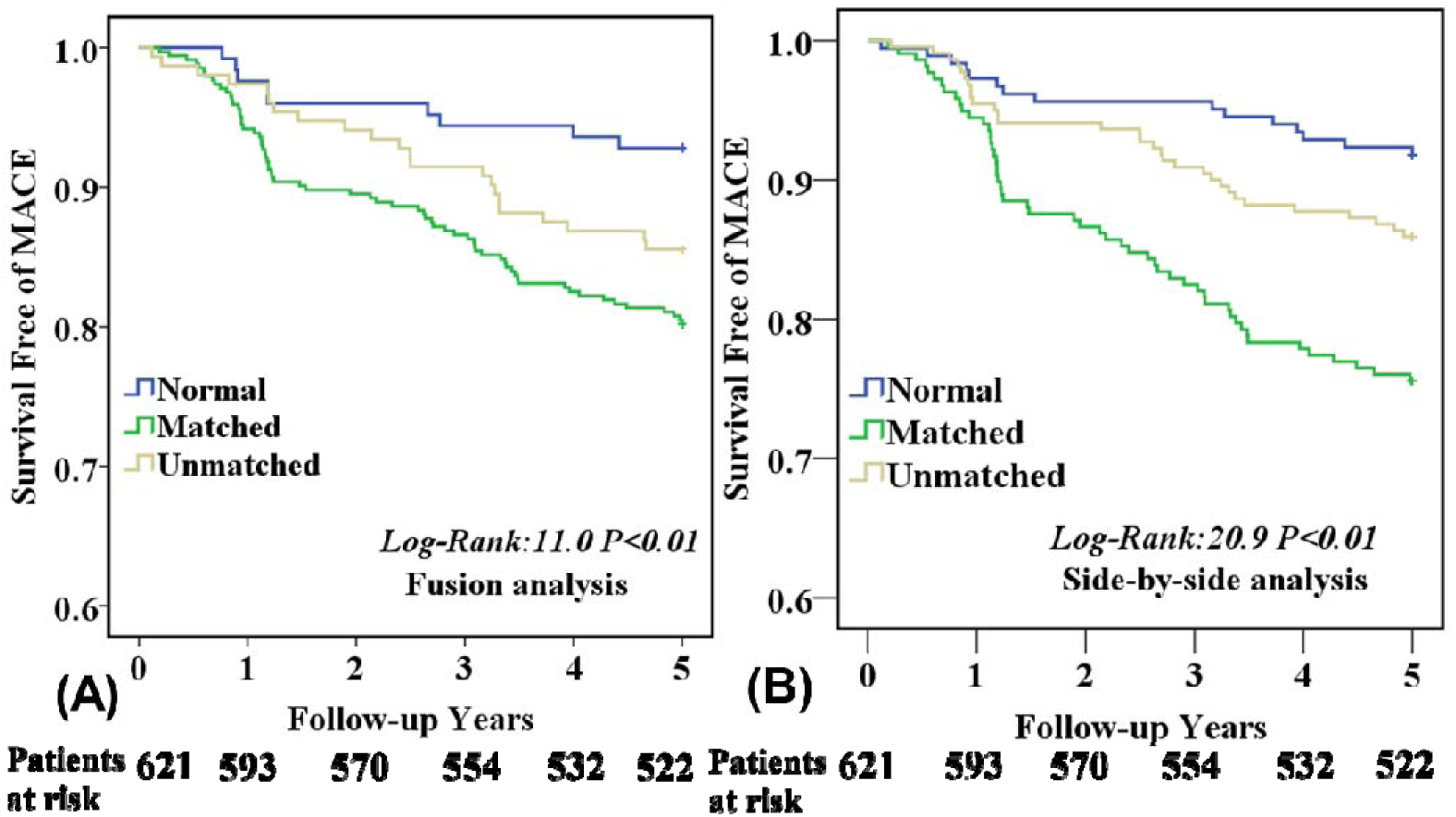
Kaplan-Meier survival curves showing the prognostic value of the fusion and side-by-side analysis on patients with known or suspected CAD (N=621.

### Influence of Revascularization on Outcomes

Patients with at least one vessel stenosis above 50% were observed in 366 out of 621 patients. Fig.3 A shows that patients who received revascularization concordant with the guidance of the fusion were associated with significantly better survival when compared to those who received revascularization when findings were discordant on fusion analysis (16.8% vs 27.0%, P<0.05). In contrast, there was no difference in survival with revascularization in the groups with side-by-side analysis, regardless of the concordance of revascularization (19.5% VS 24.2%, P>0.05). Additionally, the beneficial effect of revascularization was mainly in intermediate stenosis patients whose treatment was concordant with the guidance of the fusion (concordant: 10.8% vs discordant: 26.5%, P<0.01), as shown in Figure 3B. There was no significant difference in the MACE-free survival between patients with treatment concordance and discordance using either fusion or side-by-side analysis, regardless of whether they had one or multiple vessel disease, as depicted in Figure 3C and 3D.

**Figure 3.**
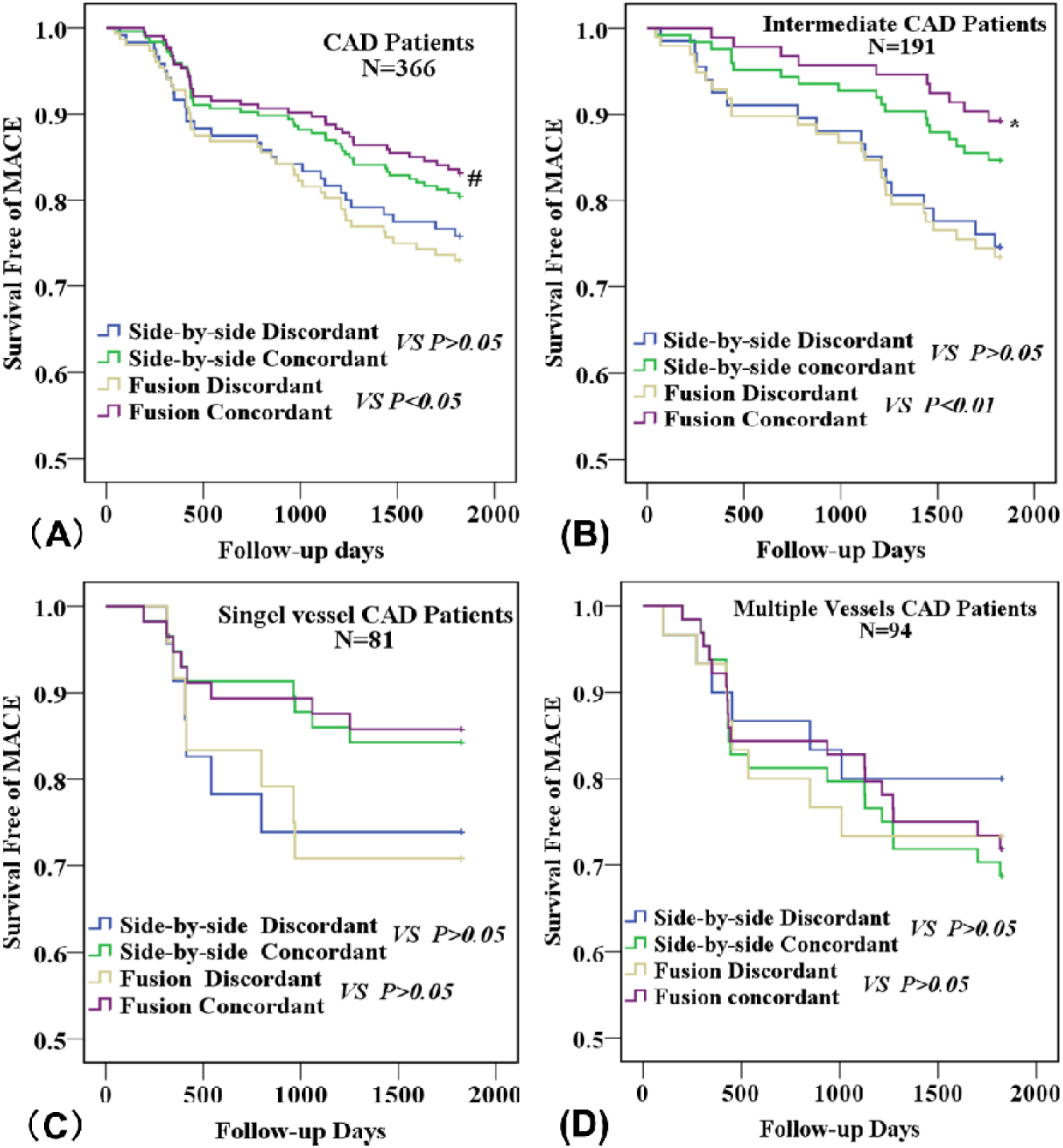
The Kaplan-Meier curves for treatments concordant with the fusion and side-by-side analysis. (A), in the total CAD patients; (B), in sub-group patients with intermediate stenosis; (C), in sub-group patients with single vessel stenosis; (D), in sub-group patients with multiple vessels stenosis (D). ***#: P<0.05; *: P<0.01*.**

Furthermore, revascularization concordant with the guidance of the fusion was a significant protective factor against MACE among CAD patients (HR:0.58, CI:0.37-0.91; P<0.01), but not the revascularization concordant with side-by-side analysis (HR:0.77, CI:0.48-1.22; P=0.27) (Fig.4 A). The protective effect of revascularization concordant with the fusion against MACE remained unchanged after the multivariate Cox regression analysis (HR:0.48, CI:0.28-0.83; P<0.01) (Fig.4 B).

**Figure 4.**
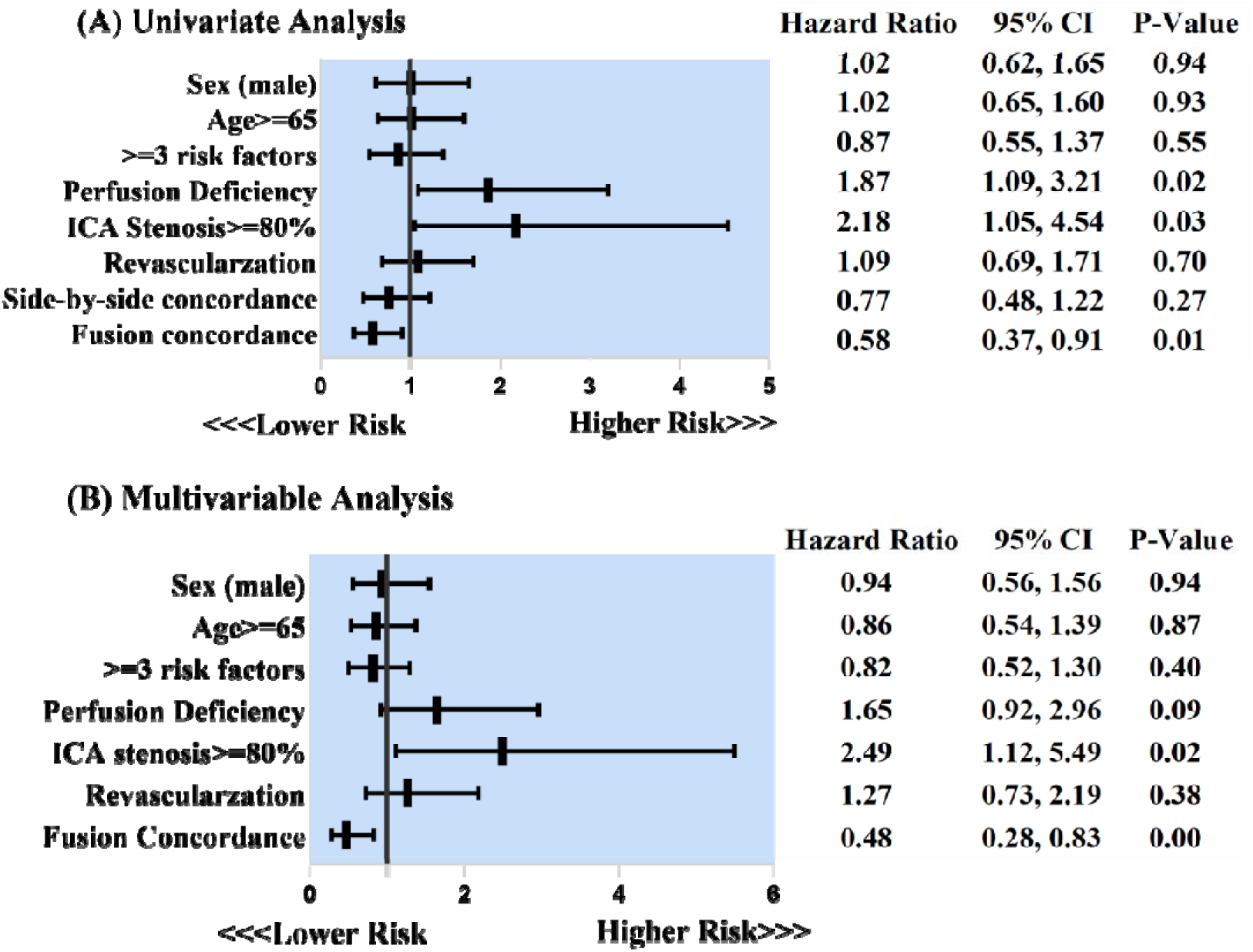
Association between covariates and outcomes in the total CAD patients (N=366). CI: confidence interval.

The unaltered protective effect of fusion-guided revascularization against MACE was also confirmed in the multivariate Cox regression analysis, both in the SDS >=10% (HR:0.40, CI:0.20-0.80;P=0.01) (Table 4 A) and SDS<10% groups (HR:0.48, CI:0.24-0.95; P=0.04)(Table 4 B).

**Table 4.** Predictors of events at univariate and multivariate analyses in patients A. SD%≥10% (N=338)

| Predictors | MACE |  |  |  |
| --- | --- | --- | --- | --- |
|  | Univariate HR<br>(95% CI) | P-value | Multivariate HR<br>(95% CI) | P-value |
| **SD% ≥ 10%** |  |  |  |  |
| Male gender | 1.09 (0.56-2.16) | 0.78 | 0.99 (0.55-1.75) | 0.96 |
| Age (≥65) | 1.62 (0.83-3.16) | 0.15 | 1.69 (0.94-2.96) | 0.06 |
| ≥3 risk factors | 1.03 (0.53-2.00) | 0.91 | 0.98 (0.58-1.72) | 0.94 |
| Fusion concordance | 0.28 (0.14-0.56) | **<0.001** | 0.40 (0.20-0.80) | **0.01** |
| Side-by-side<br>concordance | 0.54(0.28-1.05) | 0.07 | 0.87 (0.46-1.65) | 0.67 |
| Stenosis>50% | 4.79 (1.69-13.58) | **<0.001** | 2.42 (1.11-5.28) | **0.03** |
| Revascularization | 1.76 (0.90-3.44) | 0.09 | 1.39 (0.70-2.75) | 0.35 |

B.SD%<10% (N=283)
| Predictors<br>(SD%<10%) | MACE |  |  |  |
| --- | --- | --- | --- | --- |
|  | Univariate HR<br>(95% CI) | P-value | Multivariate HR<br>(95% CI) | P-value |
| Male gender | 0.94 (0.52-1.57) | 0.87 | 0.89 (0.47-1.69) | 0.72 |
| Age (≥65) | 1.09 (0.61-1.93) | 0.78 | 1.01 (0.57-1.81) | 0.96 |
| ≥3 risk factors | 0.84 (0.43-1.62) | 0.61 | 0.77 (0.38-1.49) | 0.41 |
| Fusion concordance | 0.49 (0.27-0.86) | **0.01** | 0.48 (0.24-0.95) | **0.04** |
| Side-by-side concordance | 0.79 (0.44-1.46) | 0.46 | 1.04 (0.53-2.06) | 0.91 |
| Stenosis>50% | 2.27 (1.20-4.29) | **0.01** | 1.44 (0.66-3.13) | 0.36 |
| Revascularization | 1.88 (1.04-3.39) | **0.03** | 1.95 (0.93-4.08) | 0.08 |

MACE, major adverse cardiac events; HR, hazard ratio; CI, confidence interval; NA, not applicable

### Management of patients with intermediate stenosis

Out of the 191 CAD patients with intermediate stenosis, 33 patients underwent revascularization. The lesion vessels included 141 in the LAD, 43 in the LCX, and 51 in the RCA.The revascularization procedures involved 24 LAD segments/branches, 11 LCX segments/branches, and 7 RCA segments/branches.

Fig.5 A shows that the fusion concordance group has a significantly higher lesion rate of the LCX (31.2% vs. 14.3%, P<0.01) but not in the LAD (76.1% vs. 72.6%, P>0.05) or RCA (26.9% vs. 26.6%, P>0.05), compared to the discordance group. There were no significant differences in the lesion rates of LAD (76.1% vs. 72.6%, P>0.05), LCX (16.4% vs. 25.8%, P>0.05), and RCA (26.9% vs. 26.6%, P>0.05) between patients whose treatment was concordant with the guidance of the side-by-side analysis and those whose treatment was not (Fig. 5 B).

**Figure 5.**
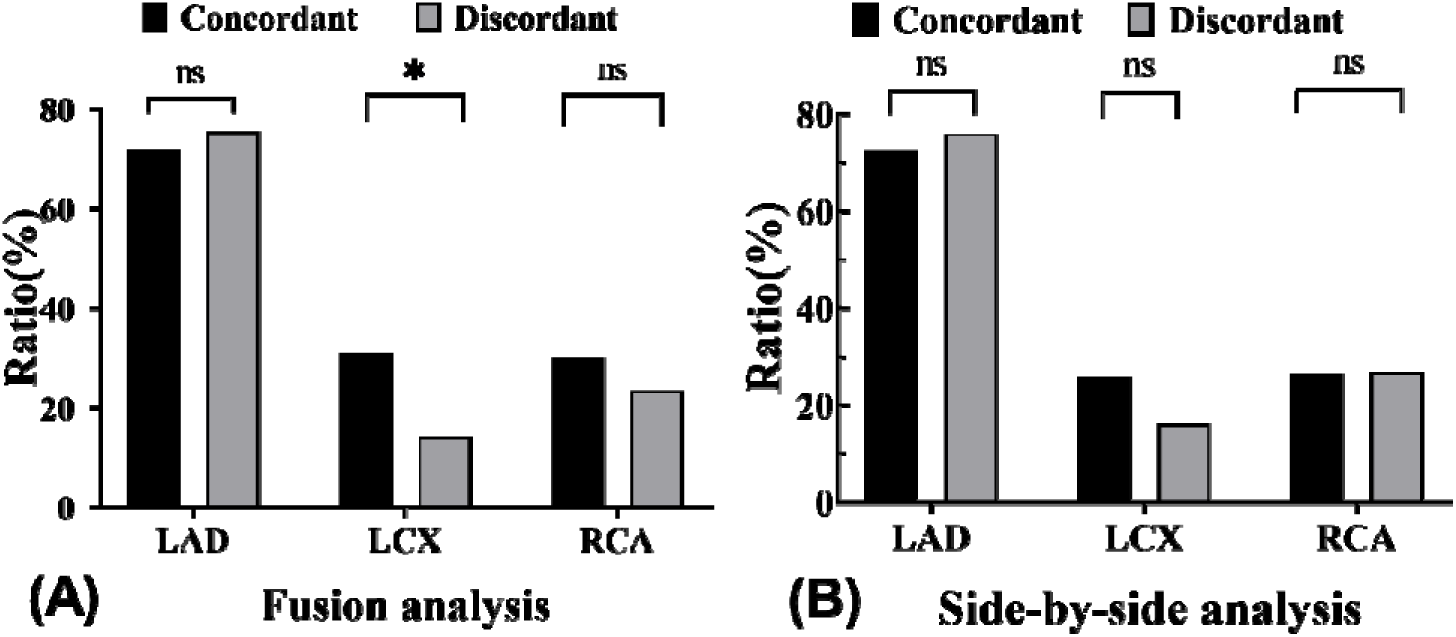
Lesion vessel ratio of the patients whose treatment was concordant/ discordant with the guidance of the fusion or side-by-side analysis. Black: treatment strategy concordant with the analysis; grey: treatment strategy discordant with the analysis; *\*: P<0.01;* ns: non-significant.

A and C show the matched patients; B and D show the unmatched patients. The yellow arrows indicate vessel stenosis, and * indicates the territory of perfusion deficit.

## Discussion

### Main Findings

In this study, we found that: 1). survival benefits after revascularization was primarily observed in the fusion concordance group compared to the discordance group in patients with known or suspected CAD, particularly in the sub-group of patients with intermediate stenosis; 2). the disparity in treatment strategies between fusion concordance and discordance was predominantly observed in the LCX, rather than in the LAD or RCA; 3). fusion concordance was identified as an independent protective factor against MACE in CAD patients.

### Benefits of the SPECT-ICA Fusion

SPECT and CTA have individually shown effectiveness in risk stratification for CAD, providing valuable information for diagnosis and evaluation [12–14]. However, the fusion of SPECT and CTA imaging can further enhance their advantages by combining and allocating perfusion deficit to the corresponding coronary arteries, thereby offering additional clinical value. The study by Pazhenkottil et al. highlighted the effectiveness of SPECT-CTA hybrid imaging in risk stratification, showing higher rates of MACE (6.0%) and revascularization (41%) in patients with matched findings compared to unmatched or normal findings [6]. Our SPECT-ICA fusion study similarly found that patients with a matched finding had the highest rate of MACE in both the fusion (19.8%) and side-by-side (24.4%) groups, along with a higher revascularization rate. Although our study did not utilize two independent cohorts, the findings still support the conclusion that fusion analysis exhibits excellent risk stratification ability in patients who have known or are suspected to have CAD. This is particularly valuable for patients who have stable CAD but don’t have symptoms, as fusion analysis can serve as a gatekeeper to prevent unnecessary revascularization[15]. Figure 6 shows examples of matched and unmatched conditions, highlighting the role of fusion imaging in resolving inconclusive results from standalone SPECT and ICA procedures. In our opinion, SPECT-ICA fusion imaging can offer improved diagnostic accuracy compared to SPECT-CTA, by elimination of motion artifacts and the reduction of severe calcification influence of CTA, which needs further study.

**Figure 6.**
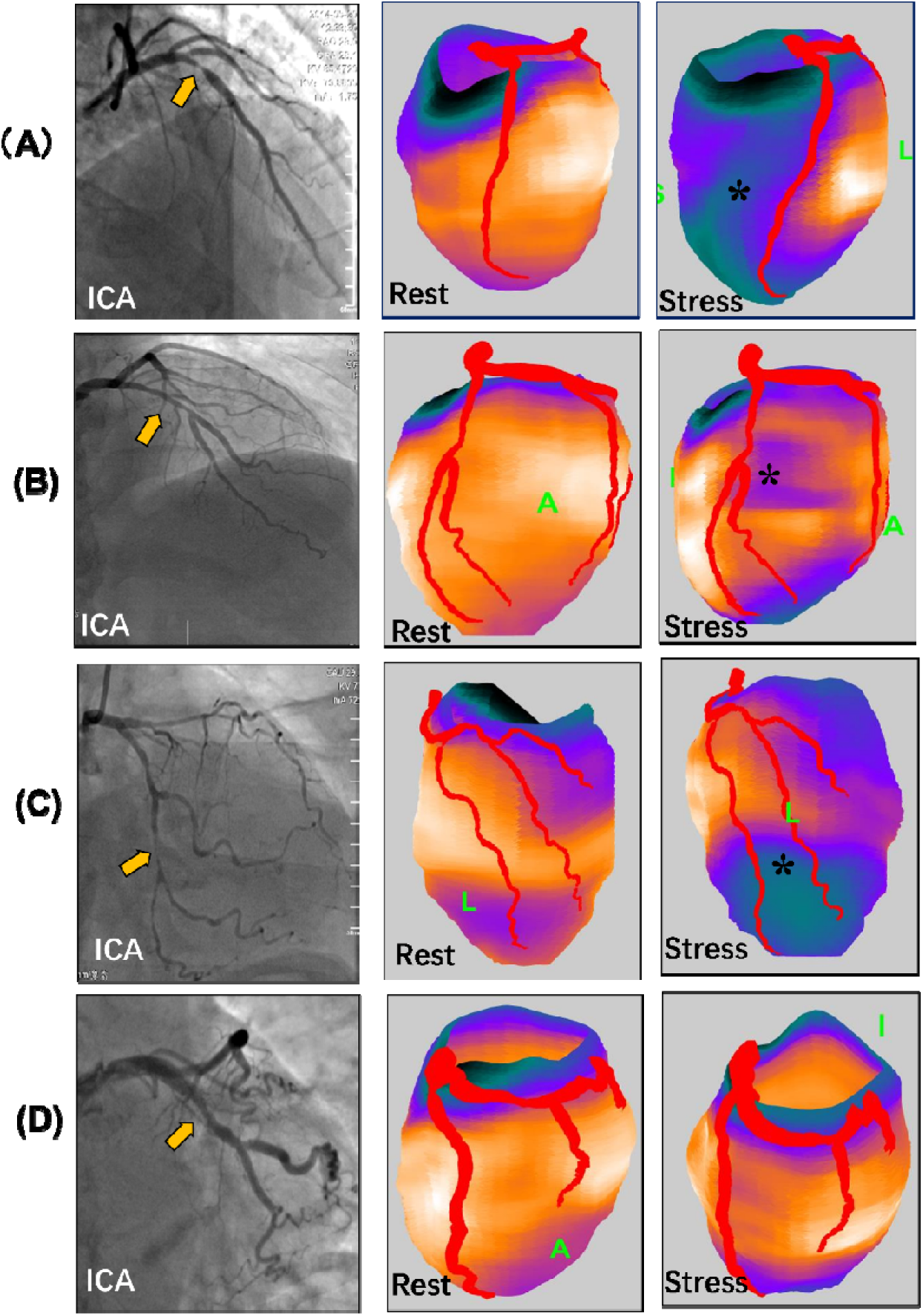
SPECT-ICA fusion showing both perfusion deficit and the corresponding vessel stenosis.

Although numerous studies have demonstrated the potential role of SPECT-CTA as a gatekeeper before revascularization [16, 17], our work offers additional insights into identifying which subgroup of patients based on the stenosis level are more likely to benefit most from integrated analysis of fused image. We observed a significantly lower MACE rate in the fusion concordance group compared to the discordance group in the 366 CAD patients (16.8% vs. 26.9%, P<0.05). However, there was no significant difference in the MACE rate between the side-by-side concordance and discordance groups (19.5% vs. 24.2%, P>0.05). After adjusted for clinical characteristics, the multiple Cox regression analysis demonstrated that fusion concordance remained an independent protective factor against MACE while side-by-side concordance did not. These results were similar to the study by Benz et al., which shows that revascularization benefits were primarily observed in the matched group. The multivariate analysis shows the early revascularization appeared to be a protective factor of MACE among patients with a matched finding [18]. However, our study defines the concordance population as patients in the matched group who received revascularization and those in the normal group who did not. From our perspective, this classification of “concordance” allows for the inclusion of more eligible patients who are more likely to benefit from integrated analysis, providing a better reflection of actual clinical practice. Moreover, when we compare the revascularization benefits in the sub-groups of CAD patients, the MACE difference only exists in the intermediate stenosis group rather than in single and multiple-vessel CAD patients. From the above results, the benefit of fusion concordance in the CAD population was mainly observed in the population with intermediated stenosis. Our finding is similar to the study by Jeroen et al., which used invasive Fractional Flow Reserve (FFR)<0.8 as a reference for significant CAD and found that the advantage of SPECT/CTA hybrid imaging was best reflected in a group of patients with intermediate to high-risk CAD [19].

Most existing studies compare hybrid imaging over standalone imaging modalities, and few studies compare the hybrid image with the side-by-side interpretation by visual fusion from the two modalities [20]. In our study, the lack of a significant difference in MACE between the side-by-side concordance and discordance groups (P>0.05) highlights the crucial role of fusion imaging in guiding the treatment of patients with intermediate stenosis. This may be caused by the practice that these side-by-side interpretations are typically reviewed through a subjective, visual integration of the results from SPECT and ICA.

To investigate the reasons for survival differences, we analyzed the lesion vessel ratio in the fusion concordance and discordance groups. The lesion vessel ratio showed no difference between the LAD and RCA, except for the LCX. In our previous study involving 36 CAD patients, we observed a significant decrease in equivocal coronary segments from 29 to 9 after the fusion analysis was performed [8]. The reclassification of vessels by the fusion analysis was primarily observed in the LCX vessels, highlighting the important role of SPECT-ICA fusion in identifying lesions in the LCX (Fig.6). These findings align with the study by Liga et al, which reported a high segmental reclassification rate (49%) in the standard LCX after the SPECT-CTA analysis[21].

### SPECT-ICA Fusion for Decision-Making of Revascularization in the Post-stent Era

The revascularization should be based on ischemia or anatomic stenosis remains controversial. The ISCHEMIA trial in patients with stable CAD and moderate to severe ischemia showed that an initial invasive strategy did not reduce the risk of all-cause mortality in any ischemia or CAD subgroup and concluded that ischemia severity was not associated with increased risk after adjustment for CAD severity [22, 23]. In this study, the independent protective value of revascularization was only observed in the univariate analysis but not multivariate analysis. However, the Fractional Flow Reserve versus Angiography for Multivessel Evaluation (FAME) serial studies revealed that higher risk populations could obtain clinical benefits from ischemia-guided revascularization [24]. Monika et al. also stated that patients should be referred for revascularization in significant stress ischemia (SD%>10%) [9]. Our study observed that revascularization concordant with the fusion was identified as an independent protective factor against MACE in both SD% _≥_ 10% and SD%**<**10% patients. However, no such effect was observed in the revascularization group concordant with the side-by-side analysis (Table 4).

In the post-stent era, the development of intravascular imaging and hemodynamic testing could supply more evidence prior to revascularization. However, the additional procedural steps and increased radiation exposure time have hindered their widespread use. It is reported that only 30% of decisions rely on intravascular diagnostic tools like FFR, intravascular ultrasound (IVUS), and optical coherence tomography (OCT), while the remaining 70% use angiographic appearance [25]. Hence, there is a compelling demand for a diagnostic tool that can offer comprehensive insights into both anatomical stenosis and ischemic burden concurrently without extra steps. Fusion imaging, which could allow an accurate assignment of myocardial perfusion regions to the corresponding vessels, had been studied a long time before [26].

Current guidelines recommend that patients with an intermediate/high pretest probability for stable CAD undergo CTA. In contrast, patients with a high pretest probability, inconclusive noninvasive testing, or refractory symptoms should be referred for ICA [27]. Accordingly, the SPECT-ICA fusion has promising clinical values. By utilizing various deep learning-based algorithms, we have been able to automatically extract left ventricles from myocardial perfusion images [28] and arterial segments from ICA frames [10, 11]. Our SPECT-ICA fusion can be completed within 5 minutes [7]. In addition to myocardial perfusion, our SPECT-ICA fusion has additional values by providing information about wall motion and thickening. This comprehensive approach allows for simultaneous evaluation of multiple parameters, enabling more accurate diagnosis and improved management of other conditions such as coronary slow flow, myocardial bridge, and microvascular disease, which will be further investigated in our future studies.

### Limitations

This is a single-center and retrospective study, which may introduce bias and limit the generalizability of the findings. The lack of a widely accepted gold standard, such as invasive FFR, makes it challenging to compare the diagnostic performance of SPECT-ICA fusion. However, these limitations do not undermine our analysis of MACE events. Other factors such as fixed perfusion deficits, reversible abnormalities, and myocardial infarction with non-obstructive coronary arteries (MINOCA) could confound the effects of revascularization. Additionally, the extent of ischemia was not quantified, which could potentially impact the diagnostic accuracy of the study.

## Conclusions

3D fusion prior to coronary revascularization can guide the treatment to improve outcomes among patients with known or suspected CAD, particularly those with intermediate stenosis.

## Data Availability

Data available on request from the authors

## Acknowledgment

This research was supported by a seed grant from Michigan Technological University Health Research Institute (PI: Weihua Zhou), and a grant from the National Nature Science Foundation of China (PI: Cheng Wang, 82100338).

## References

1. Maron DJ, Hochman JS, Reynolds HR, Bangalore S, O’Brien SM, Boden WE, et al. Initial Invasive or Conservative Strategy for Stable Coronary Disease. N Engl J Med. 2020;382(15):1395–1407. 10.1056/NEJMoa1915922.

2. Tonino PA, De Bruyne B, Pijls NH, Siebert U, Ikeno F, Van’ TVM, et al. Fractional flow reserve versus angiography for guiding percutaneous coronary intervention. N Engl J Med. 2009;360(3):213–24. 10.1056/NEJMoa0807611.

3. Bourque JM, Beller GA. Stress myocardial perfusion imaging for assessing prognosis: an update. Jacc Cardiovasc Imaging. 2011;4(12):1305–19. 10.1016/j.jcmg.2011.10.003.

4. Shiraishi S, Sakamoto F, Tsuda N, Yoshida M, Tomiguchi S, Utsunomiya D, et al. Prediction of left main or 3-vessel disease using myocardial perfusion reserve on dynamic thallium-201 single-photon emission computed tomography with a semiconductor gamma camera. Circ J. 2015;79(3):623–31. 10.1253/circj.CJ-14-0932.

5. Gaemperli O, Saraste A, Knuuti J. Cardiac hybrid imaging. Eur Heart J Cardiovasc Imaging. 2012;13(1):51–60. 10.1093/ejechocard/jer240.

6. Pazhenkottil AP, Benz DC, Gräni C, Madsen MA, Mikulicic F, von Felten E, et al. Hybrid SPECT Perfusion Imaging and Coronary CT Angiography: Long-term Prognostic Value for Cardiovascular Outcomes. Radiology. 2018;288(3):694–702. 10.1148/radiol.2018171303.

7. Tang H, Bober RR, Zhao C, Zhang C, Zhu H, He Z, et al. 3D fusion between fluoroscopy angiograms and SPECT myocardial perfusion images to guide percutaneous coronary intervention. J Nucl Cardiol. 2021. 10.1007/s12350-021-02574-1.

8. Xu Z, Tang H, Malhotra S, Dong M, Zhao C, Ye Z, et al. Three-dimensional Fusion of Myocardial Perfusion SPECT and Invasive Coronary Angiography Guides Coronary Revascularization. J Nucl Cardiol. 2022. 10.1007/s12350-022-02907-8.

9. Czaja M, Wygoda Z, Duszanska A, Szczerba D, Glowacki J, Gasior M, et al. Interpreting myocardial perfusion scintigraphy using single-photon emission computed tomography. Part 1. Kardiochir Torakochirurgia Pol. 2017;14(3):192–199. 10.5114/kitp.2017.70534.

10. Zhao C, Vij A, Malhotra S, Tang J, Tang H, Pienta D, et al. Automatic extraction and stenosis evaluation of coronary arteries in invasive coronary angiograms. Comput Biol Med. 2021;136:104667. 10.1016/j.compbiomed.2021.104667.

11. Zhao C, Xu Z, Jiang J, Esposito M, Pienta D, Hung G, et al. AGMN: Association graph-based graph matching network for coronary artery semantic labeling on invasive coronary angiograms. Pattern Recognit. 2023;143:109789. 10.1016/j.patcog.2023.109789.

12. Douglas PS, Hoffmann U, Patel MR, Mark DB, Al-Khalidi HR, Cavanaugh B, et al. Outcomes of anatomical versus functional testing for coronary artery disease. The New England Journal of Medicine. 2015;372(14):1291–300. 10.1056/NEJMoa1415516.

13. Perrone-Filardi P, Achenbach S, Mohlenkamp S, Reiner Z, Sambuceti G, Schuijf JD, et al. Cardiac computed tomography and myocardial perfusion scintigraphy for risk stratification in asymptomatic individuals without known cardiovascular disease: a position statement of the Working Group on Nuclear Cardiology and Cardiac CT of the European Society of Cardiology. Eur Heart J. 2011;32(16):1986–93, 1993a, 1993b. 10.1093/eurheartj/ehq235.

14. Gimelli A, Pugliese NR, Buechel RR, Coceani M, Clemente A, Kaufmann PA, et al. Myocardial perfusion scintigraphy for risk stratification of patients with coronary artery disease: the AMICO registry. Eur Heart J Cardiovasc Imaging. 2020. 10.1093/ehjci/jeaa298.

15. Giannopoulos AA, Gaemperli O. Hybrid Imaging in Ischemic Heart Disease. Rev Esp Cardiol (Engl Ed). 2018;71(5):382–390. 10.1016/j.rec.2017.11.023.

16. Pazhenkottil AP, Nkoulou RN, Ghadri JR, Herzog BA, Buechel RR, Küest SM, et al. Prognostic value of cardiac hybrid imaging integrating single-photon emission computed tomography with coronary computed tomography angiography. Eur Heart J. 2011;32(12):1465–71. 10.1093/eurheartj/ehr047.

17. Pazhenkottil AP, Nkoulou RN, Ghadri JR, Herzog BA, Kuest SM, Husmann L, et al. Impact of cardiac hybrid single-photon emission computed tomography/computed tomography imaging on choice of treatment strategy in coronary artery disease. Eur Heart J. 2011;32(22):2824–9. 10.1093/eurheartj/ehr232.

18. Benz DC, Gaemperli L, Gräni C, von Felten E, Giannopoulos AA, Messerli M, et al. Impact of cardiac hybrid imaging-guided patient management on clinical long-term outcome. Int J Cardiol. 2018;261:218–222. 10.1016/j.ijcard.2018.01.118.

19. Schaap J, Kauling RM, Boekholdt SM, Nieman K, Meijboom WB, Post MC, et al. Incremental diagnostic accuracy of hybrid SPECT/CT coronary angiography in a population with an intermediate to high pre-test likelihood of coronary artery disease. Eur Heart J Cardiovasc Imaging. 2013;14(7):642–9. 10.1093/ehjci/jes303.

20. Kirisli HA, Gupta V, Shahzad R, Al YI, Dharampal A, Geuns RJ, et al. Additional diagnostic value of integrated analysis of cardiac CTA and SPECT MPI using the SMARTVis system in patients with suspected coronary artery disease. J Nucl Med. 2014;55(1):50–7. 10.2967/jnumed.113.119842.

21. Liga R, Vontobel J, Rovai D, Marinelli M, Caselli C, Pietila M, et al. Multicentre multi-device hybrid imaging study of coronary artery disease: results from the EValuation of INtegrated Cardiac Imaging for the Detection and Characterization of Ischaemic Heart Disease (EVINCI) hybrid imaging population. Eur Heart J Cardiovasc Imaging. 2016;17(9):951–60. 10.1093/ehjci/jew038.

22. Reynolds HR, Shaw LJ, Min JK, Page CB, Berman DS, Chaitman BR, et al. Outcomes in the ISCHEMIA Trial Based on Coronary Artery Disease and Ischemia Severity. Circulation. 2021;144(13):1024–1038. 10.1161/CIRCULATIONAHA.120.049755.

23. Hochman JS, Anthopolos R, Reynolds HR, Bangalore S, Xu Y, O’Brien SM, et al. Survival After Invasive or Conservative Management of Stable Coronary Disease. Circulation. 2023;147(1):8–19. 10.1161/CIRCULATIONAHA.122.062714.

24. Xaplanteris P, Fournier S, Pijls N, Fearon WF, Barbato E, Tonino P, et al. Five-Year Outcomes with PCI Guided by Fractional Flow Reserve. N Engl J Med. 2018;379(3):250–259. 10.1056/NEJMoa1803538.

25. Cho H, Lee JG, Kang SJ, Kim WJ, Choi SY, Ko J, et al. Angiography-Based Machine Learning for Predicting Fractional Flow Reserve in Intermediate Coronary Artery Lesions. J Am Heart Assoc. 2019;8(4):e011685. 10.1161/JAHA.118.011685.

26. Peifer JW, Ezquerra NF, Cooke CD, Mullick R, Klein L, Hyche ME, et al. Visualization of multimodality cardiac imagery. Ieee Trans Biomed Eng. 1990;37(8):744–56. 10.1109/10.102790.

27. Gulati M, Levy PD, Mukherjee D, Amsterdam E, Bhatt DL, Birtcher KK, et al. 2021 AHA/ACC/ASE/CHEST/SAEM/SCCT/SCMR Guideline for the Evaluation and Diagnosis of Chest Pain: A Report of the American College of Cardiology/American Heart Association Joint Committee on Clinical Practice Guidelines. Circulation. 2021;144(22):e368–e454. 10.1161/CIR.0000000000001029.

28. Zhu F, Li L, Zhao J, Zhao C, Tang S, Nan J, et al. A new method incorporating deep learning with shape priors for left ventricular segmentation in myocardial perfusion SPECT images. Comput Biol Med. 2023;160:106954. 10.1016/j.compbiomed.2023.106954.

